# Ultrasound Detection of Early Callus Formation in Proximal Humerus Fractures: Protocol for a Pilot and Prospective Cohort Study

**DOI:** 10.64898/2026.07.20.26358520

**Authors:** Benjamin Blackman, Niall Fahey, Stephen Dolan, Michael Kevin O’Reilly, John Tristan Cassidy

## Abstract

**Introduction:** Proximal humerus fractures account for approximately 5–6% of all adult fractures and are primarily managed nonoperatively. Healing is conventionally monitored with radiographs, with radiopaque callus formation indicating healing. Visible radiographic callus appears weeks after biological union begins. Ultrasound provides a dynamic, radiation-free, and cost-effective method that can detect early callus formation before x-ray visibility. Although ultrasound has demonstrated utility for fracture healing in the clavicle and humeral shaft, its role in proximal humerus fractures remains unclear.

**Methods:** This single-centre prospective study will be conducted in two phases. The pilot phase will measure inter-rater reliability for ultrasound detection of early callus formation at 2 and 4 weeks post-injury. Ten patients with proximal humerus fractures treated nonoperatively will undergo standardized anterior and lateral scans. Each patient will generate four saved images (short- and long-axis views), producing forty anonymized images independently reviewed by two raters. The prospective cohort phase will recruit approximately thirty additional patients.

**Results:** Reliability will be quantified using Cohen’s kappa. A power calculation will be performed after pilot analysis. Results from the prospective cohort phase will help determine the association and predictive value of early ultrasound-detected bridging callus for radiographic and clinical union at three and six months. Patient reported outcome measures will be assessed using the Quick Disabilities of Arm, Shoulder and Hand (QuickDASH) questionnaire.

**Discussion:** This study will develop and validate a standardized ultrasound protocol for assessing early fracture healing in proximal humerus fractures. By establishing both inter-rater reliability and predictive value, the findings may support ultrasound as a reproducible, radiation-free adjunct to conventional imaging and enable earlier identification of union status.

## INTRODUCTION

Proximal humerus fractures comprise 5–6% of all adult fractures, most often due to low-energy falls in older adults with osteoporotic bone [1]. These injuries can lead to long-term functional impairment and reduced quality of life [2]. Stable, extra-articular or minimally displaced fractures are typically treated non-operatively with sling immobilization and early physiotherapy [3].

Fracture healing is a multistage process involving hematoma formation, callus development, and bone remodeling [4,5]. Progress toward union is assessed through serial radiographs, where visible callus is a key marker of healing [6]. However, radiographic callus often appears weeks after biological union begins.

Ultrasound is an alternative imaging modality for various musculoskeletal conditions [7,8]. Ultrasound offers a non-invasive, radiation-free, and dynamic method to visualize early fracture healing, capable of detecting hypoechoic fibrocartilaginous and hyperechoic osseous callus before radiographic evidence [9–11]. Prior studies demonstrated that ultrasound-detected bridging callus predicts union in clavicle and humeral shaft fractures [12–15]. Nevertheless, its role in proximal humerus fractures is poorly defined, and user dependence remains a limitation [16].

A recent review reported that ultrasound may predict healing capacity [17]. Despite this potential, its diagnostic use in fracture management is not yet routinely practiced [17]. Our pilot study aims to standardize an ultrasound protocol for proximal humerus fractures and assess inter-rater reliability in callus detection. The subsequent cohort phase will explore whether early sonographic bridging callus correlates with radiographic and clinical union.

## METHODS

### Study Design

This is a single-centre prospective cohort study conducted in the Department of Trauma and Orthopaedics at University Hospital Limerick, Ireland. The study includes a pilot reliability phase followed by a longitudinal cohort phase. Ethical approval was obtained from the Health Service Executive Research Ethics Committee (REC Ref: 0171).

### Study Population

Patients presenting to the fracture clinic at University Hospital Limerick with proximal humerus fractures treated nonoperatively will be eligible for inclusion.

### Inclusion criteria

1. Are ≥ 18 years of age
2. Radiographically confirmed proximal humerus fracture
3. Are managed nonoperatively at our institution
4. Present to fracture clinic within 14 days of injury

### Exclusion criteria

1. Proximal humerus fractures treated surgically
2. Pathological fractures
3. Polytrauma or multiple fractures of the ipsilateral upper limb that would interfere with ultrasound assessment or clinical scoring
4. Previous major surgery, fracture, or advanced arthritis of the ipsilateral shoulder
5. Inability to comply with follow-up

### Phase 1: Training and Pilot Reliability Study

From April 2025 to December 2025, ten patients will be enrolled in a two-stage process. Sample size for the pilot phase is consistent with prior reliability studies evaluating ultrasound for fracture healing [13]. A musculoskeletal sonographer, a consultant radiologist and fellowship-trained orthopaedic shoulder surgeon will perform initial ultrasound examinations on ten patients with proximal humerus fractures at 2 and 4 weeks following injury to achieve consensus on the sonographic appearance of callus. The cortical bone will be evaluated for periosteal elevation, cortical discontinuity, step-off deformity with one or more hyperechoic reflections, and the “double-line sign,” as well as the presence of hypoechoic fibrocartilaginous or hyperechoic early osseous callus, which together assess both fracture disruption and early healing [9,13,14,17,18]. Standard follow-up radiographs will be reviewed concurrently to confirm fracture location and stage of healing. Each patient will have four standardized scans: anterior and lateral planes in both short- and long-axis. There will be forty images generated in total. To minimize bias, images showing both callus and no callus will be included. Each image will be labeled with patient identifier, date, and view, then anonymized and randomized by a researcher not involved in scanning.

A consultant radiologist and orthopaedic surgeon will independently assess all anonymized images for the presence or absence of early callus formation. Inter-rater agreement will be quantified using Cohen’s κ [19]. Accuracy will be compared against the consensus assessment during training and radiographic reference where appropriate.

### Phase 2: Longitudinal Cohort Study

Between January 2026 and Dec 2026, we aim to enroll approximately thirty additional patients for longitudinal assessment. Ultrasound will be performed at 2 and 4 weeks post-injury, with radiographs obtained per standard care at our institution (2 weeks, 4 weeks, 3 months, 6 months). Early sonographic bridging callus will be correlated with eventual radiographic and clinical union. A formal power calculation for this phase will be completed once pilot reliability data yield variance estimates. The protocol flow diagram can be seen in Figure 1.

**Figure 1.**
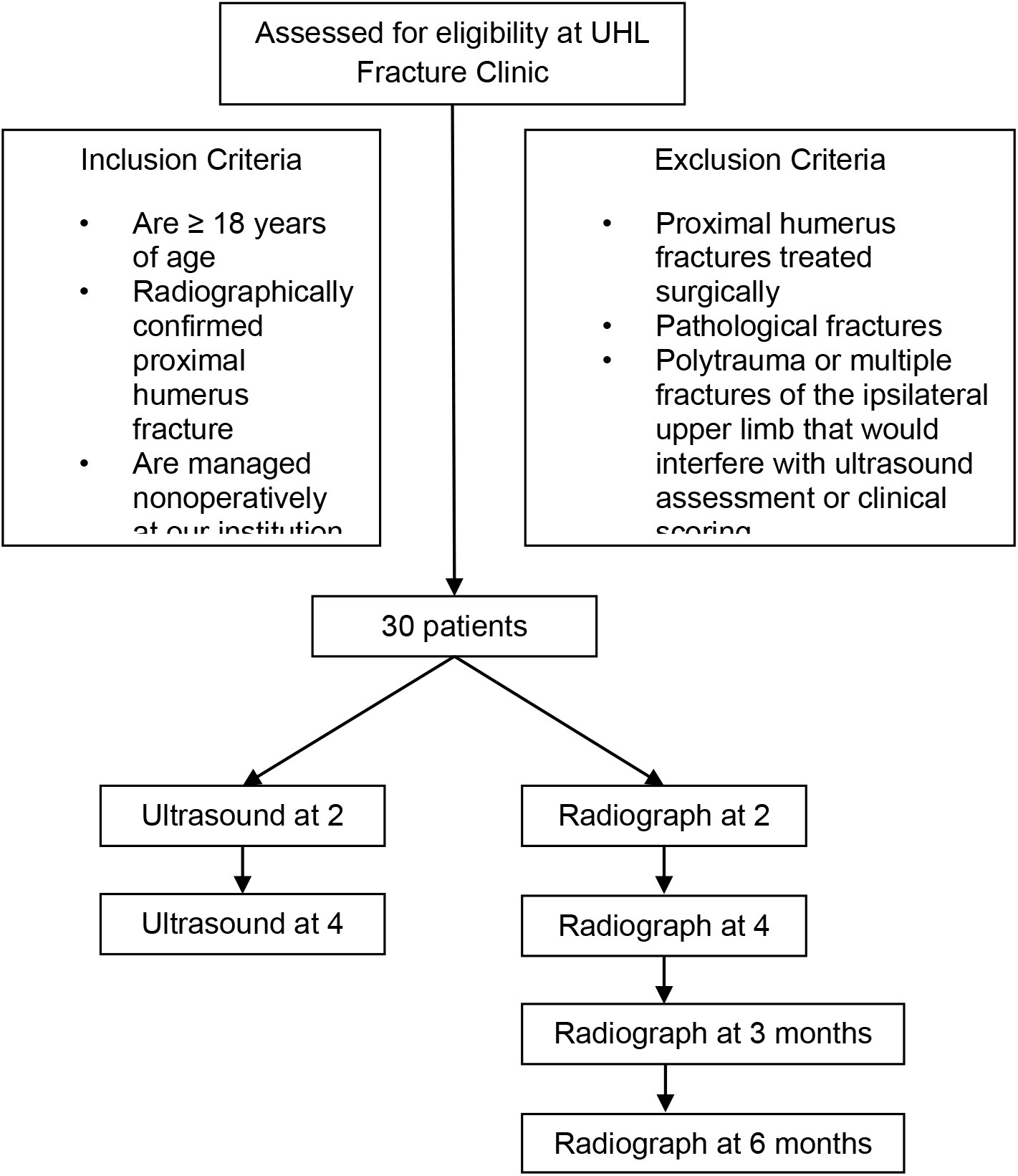
Flow Diagram for the prospective cohort study

### Ultrasound Technique

All scans will be performed using a high-frequency (6–15 MHz) linear transducer calibrated to a 3 cm depth. Ultrasounds will follow a standardized superficial musculoskeletal protocol.

### Patient positioning

Patients will be seated with the affected arm slightly abducted and resting on the ipsilateral thigh with the palm supinated.

### Anterior approach

The probe is placed at the level of the deltoid tuberosity and scanned proximally along the long head of the biceps in short axis to visualize the surgical neck, humeral head, and bicipital groove. Suspected callus is assessed with Doppler to evaluate vascularity. The sequence is repeated in long axis, and both still and loop images are saved.

### Lateral approach

The probe is placed laterally at the level of the deltoid tuberosity and scanned proximally toward the acromion. The greater tuberosity and humeral head are examined for hypoechoic fibrocartilaginous or hyperechoic osseous callus. Doppler is again used for vascular assessment, and loops and stills are captured in both short- and long-axis views.

### Ultrasound Interpretation

Scans will be classified into three categories based on sonographic appearance [9,11,14,17]. Fibrocartilaginous callus will be defined as hypoechoic tissue bridging the fracture gap. Nonbridging callus will refer to localized hyperechoic foci that are not continuous across the cortical surface. Bridging callus will be identified as a continuous hyperechoic signal with cortical bone–like echogenicity spanning the fracture ends. All still images and loop recordings will be archived under coded identifiers and securely stored within an institutional research database.

### Outcome Measures

The primary outcome of the pilot phase is the inter-rater reliability of ultrasound detection of early callus formation at 2 and 4 weeks, quantified using Cohen’s κ. In the prospective cohort phase, the primary outcome is the association and predictive value of early ultrasound-detected callus formation for subsequent radiographic and clinical union at three and six months. Patient reported outcome measures will be assessed using the Quick Disabilities of the Arm, Shoulder and Hand (QuickDASH) questionnaire [20,21]. Secondary outcomes across both phases include the accuracy of ultrasound compared with radiographs in identifying early union and the timing of bridging callus appearance on ultrasound relative to radiographic evidence of healing.

### Statistical Analysis

Descriptive statistics will summarize demographic variables. Inter-rater reliability will be calculated using Cohen’s κ. κ values will be interpreted as: 0.00–0.20 slight, 0.21–0.40 fair, 0.41–0.60 moderate, 0.61–0.80 substantial, and 0.81–1.00 almost perfect agreement [19]. Correlations between ultrasound and radiographic union will be analyzed using Pearson or Spearman coefficients as appropriate. Logistic regression will assess the predictive validity of sonographic callus for union, adjusting for age, sex, smoking, and displacement. Statistical significance will be set at *P* < 0.05.

### Data Management

All images and clinical data will be deidentified and stored on a secure hospital server compliant with General Data Protection Regulation (GDPR). Only the study investigators will have access.

### Ethics and Dissemination

Ethical approval was granted (REC Ref: 0171). Written informed consent will be obtained from all participants. Results will be disseminated through peer-reviewed journals and conference presentations.

## DISCUSSION

This study will be the first to systematically evaluate ultrasound as a tool for early detection of callus formation in proximal humerus fractures. Previous ultrasound work in long bone fractures demonstrates that sonographic callus can be detected well before radiographic visibility and can predict healing outcomes. In a prospective cohort of 112 displaced midshaft clavicle fractures, sonographic bridging callus at six weeks was associated with a 98.6 percent union rate, while its absence carried a markedly higher nonunion risk, and accuracy improved further when combined with clinical risk factors [12]. In humeral shaft fractures, ultrasound has been shown to reliably detect early healing and help identify patients at increased risk of nonunion [13]. Broader review evidence suggests that modern ultrasound processing and emerging 3D techniques may further improve diagnostic accuracy and support early recognition of impaired healing in trauma settings [15]. In surgically treated long bone fractures, ultrasonography predicted 87.5% union and 12.5% delayed or non-union as early as 6 weeks after surgery with sensitivity and specificity above 97 percent [11]. Ultrasound has also proven particularly valuable in identifying subtle or occult fractures and early callus when plain radiographs remain inconclusive in symptomatic patients [7]. Additionally, preliminary paediatric studies support ultrasound as a viable alternative to radiographs for monitoring callus formation in long bone fractures [8]. Together, this body of evidence reinforces the potential benefit of applying ultrasound to proximal humerus fractures, where earlier assessment of healing may significantly influence decision-making, rehabilitation and surveillance for delayed union.

The proposed pilot phase will generate foundational reliability data on callus detection, which will inform sample size and methodological refinement for the subsequent longitudinal cohort phase. If ultrasound can reliably identify early callus before it becomes radiographically apparent, it may enable clinicians to detect delayed or absent healing earlier in the recovery process. This could help guide more individualized rehabilitation plans, allow earlier intervention when healing is suboptimal, and potentially reduce the need for repeat radiographs. Ultrasound offers advantages of accessibility, cost-effectiveness and absence of radiation exposure, making it particularly valuable for frail or elderly patients who often require frequent imaging.

The study’s limitations include operator dependence, the modest sample size of the pilot phase and its single-centre design, which may limit generalizability.

## CONCLUSION

This protocol describes a two-phase prospective cohort study designed to assess the reliability and predictive value of ultrasound in detecting early callus formation in proximal humerus fractures. The results will determine whether sonographic bridging callus can serve as a reproducible early marker of fracture healing and help predict radiographic and clinical union. Establishing a validated, standardized ultrasound protocol could provide clinicians with a practical, radiation-free adjunct for monitoring fracture healing, facilitating earlier diagnosis of delayed union, and optimizing patient-specific rehabilitation strategies.

## Data Availability

No datasets were generated or analysed during the current study. All relevant data from this study will be made available upon study completion.

## Author Contributions

BB drafted the manuscript. NF revised manuscript and assisted with scanning protocol.SD assisted with manuscript preparation and project administration. MOR and JTC developed the scanning protocol and revised the manuscript. JTC conceived and supervised the study. All authors approved the final version and are accountable for the work

## Data Availability

Data will be made available upon reasonable request to the corresponding author.

## Ethical Approval

Approved by the Health Service Executive Research Ethics Committee (REC Ref: 0171).

## REFERENCES

1. Court-Brown CM, Caesar B. Epidemiology of adult fractures: A review. Injury. 2006;37:691– 7. 10.1016/j.injury.2006.04.130

2. de Putter CE, Selles RW, Haagsma JA, Polinder S, Panneman MJM, Hovius SER, et al. Health-related quality of life after upper extremity injuries and predictors for suboptimal outcome. Injury. 2014;45:1752–8. 10.1016/j.injury.2014.07.016

3. Baker HP, Gutbrod J, Strelzow JA, Maassen NH, Shi L. Management of Proximal Humerus Fractures in Adults—A Scoping Review. J Clin Med. 2022;11:6140. 10.3390/jcm11206140

4. Sheen JR, Mabrouk A, Garla VV. Fracture Healing Overview. StatPearls [Internet]. Treasure Island (FL): StatPearls Publishing; 2025 [cited 2025 Apr 6]. http://www.ncbi.nlm.nih.gov/books/NBK551678/. Accessed 6 Apr 2025

5. Bigham-Sadegh A, Oryan A. Basic concepts regarding fracture healing and the current options and future directions in managing bone fractures. Int Wound J. 2015;12:238–47. 10.1111/iwj.12231

6. ElHawary H, Baradaran A, Abi-Rafeh J, Vorstenbosch J, Xu L, Efanov JI. Bone Healing and Inflammation: Principles of Fracture and Repair. Semin Plast Surg. 2021;35:198–203. 10.1055/s-0041-1732334

7. Shah AB, Bhatnagar N. Ultrasound imaging in musculoskeletal injuries-What the Orthopaedic surgeon needs to know. J Clin Orthop Trauma. 2019;10:659–65. 10.1016/j.jcot.2019.05.010

8. Wawrzyk M, Sokal J, Andrzejewska E, Przewratil P. The Role of Ultrasound Imaging of Callus Formation in the Treatment of Long Bone Fractures in Children. Polish Journal of Radiology. Termedia Publishing; 2015;80:473. 10.12659/PJR.894548

9. Cocco G, Ricci V, Villani M, Delli Pizzi A, Izzi J, Mastandrea M, et al. Ultrasound imaging of bone fractures. Insights Imaging. 2022;13:189. 10.1186/s13244-022-01335-z

10. Bianchi S. Ultrasound and bone: a pictorial review. J Ultrasound. 2020;23:227–57. 10.1007/s40477-020-00477-4

11. Pinto TR, Gowda CS, Braggs AV, Mirza K, Hegde K A. The value of ultrasonography in predicting the outcomes of simple long bone fractures treated by closed intramedullary nail fixation. Chinese Journal of Traumatology [Internet]. 2023 [cited 2024 Oct 14]; 10.1016/j.cjtee.2023.11.007

12. Nicholson JA, Oliver WM, MacGillivray TJ, Robinson CM, Simpson AHRW. Sonographic bridging callus at six weeks following displaced midshaft clavicle fracture can accurately predict healing. Bone & Joint Research. Bone & Joint; 2021;10:113–21. 10.1302/2046-3758.102.BJR-2020-0341.R1

13. Oliver WM, Nicholson JA, Bell KR, Carter TH, White TO, Clement ND, et al. Ultrasound assessment of humeral shaft nonunion risk: a feasibility and proof of concept study. Eur J Orthop Surg Traumatol. 2023;34:909–18. 10.1007/s00590-023-03725-5

14. Nicholson JA, Oliver WM, LizHang J, MacGillivray T, Perks F, Robinson CM, et al. Sonographic bridging callus: An early predictor of fracture union. Injury. 2019;50:2196–202. 10.1016/j.injury.2019.09.027

15. Nicholson JA, Oliver WM, MacGillivray TJ, Robinson CM, Simpson AHRW. 3D ultrasound reconstruction of sonographic callus: a novel imaging modality for early evaluation of fracture healing. Bone & Joint Research. Bone & Joint; 2021;10:759–66. 10.1302/2046-3758.1012.BJR-2021-0250

16. Caserta MP, Bonnett SL, La Valley MC, De Meo S, Bowman AW. Ultrasound Practice Redesign to Improve Image Quality: Implementation of a Quality Control Sonographer. Journal of the American College of Radiology. 2020;17:1644–52. 10.1016/j.jacr.2020.07.015

17. Nicholson JA, Tsang STJ, MacGillivray TJ, Perks F, Simpson AHRW. What is the role of ultrasound in fracture management? Bone Joint Res. 2019;8:304–12. 10.1302/2046-3758.87.BJR-2018-0215.R2

18. Rutten MJCM, Jager GJ, de Waal malefijt MC, Blickman JG. Double line sign: a helpful sonographic sign to detect occult fractures of the proximal humerus. Eur Radiol. 2007;17:762–7. 10.1007/s00330-006-0331-1

19. Landis JR, Koch GG. The Measurement of Observer Agreement for Categorical Data. Biometrics. [Wiley, International Biometric Society]; 1977;33:159–74. 10.2307/2529310

20. Beaton DE, Wright JG, Katz JN, Group TUEC. Development of the QuickDASH: Comparison of Three Item-Reduction Approaches. JBJS. 2005;87:1038. 10.2106/JBJS.D.02060

21. Gummesson C, Ward MM, Atroshi I. The shortened disabilities of the arm, shoulder and hand questionnaire (QuickDASH): validity and reliability based on responses within the full-length DASH. BMC Musculoskelet Disord. 2006;7:44. 10.1186/1471-2474-7-44

